# Dynamic deformable attention (DDANet) for semantic segmentation

**DOI:** 10.1101/2020.08.25.20181834

**Authors:** Kumar T. Rajamani, Hanna Siebert, Mattias P. Heinrich

## Abstract

Deep learning based medical image segmentation is an important step within diagnosis, which relies strongly on capturing sufficient spatial context without requiring too complex models that are hard to train with limited labelled data. Training data is in particular scarce for segmenting infection regions of CT images of COVID-19 patients. Attention models help gather contextual information within deep networks and benefit semantic segmentation tasks. The recent criss-cross-attention module aims to approximate global self-attention while remaining memory and time efficient by separating horizontal and vertical selfsimilarity computations. However, capturing attention from all non-local locations can adversely impact the accuracy of semantic segmentation networks. We propose a new Dynamic Deformable Attention Network (DDANet) that enables a more accurate contextual information computation in a similarly efficient way. Our novel technique is based on a deformable criss-cross attention block that learns both attention coefficients and attention offsets in a continuous way. A deep segmentation network (in our case a U-Net [1]) that employs this attention mechanism is able to capture attention from pertinent non-local locations and also improves the performance on semantic segmentation tasks compared to criss-cross attention within a U-Net on a challenging COVID-19 lesion segmentation task. Our validation experiments show that the performance gain of the recursively applied dynamic deformable attention blocks comes from their ability to capture dynamic and precise (wider) attention context. Our DDANet achieves Dice scores of 73.4% and 61.3% for Ground-Glass-Opacity and Consolidation lesions for COVID-19 segmentation and improves the accuracy by 4.9% points compared to a baseline U-Net.

## I. Introduction

THE coronavirus COVID-19 pandemic is having a global impact affecting 213 countries so far. The cases world wide as reported on Worldometers [2] is about 16,482,271 as of end July 2020. Many of the countries have steadily flattened the curve by stringent social distancing measures. In the last several months of managing this pandemic globally, several screening options have become main stream from Nucleic Acid Amplification Tests (NAAT) assay tests, serological tests, and radiological imaging (X-rays, CT). Recent studies have also demonstrated that lack of taste and smell is a new indicator for this virus [3].

The gold-standard for COVID-19 diagnosis is currently using reverse-transcription polymerase chain reaction (RT-PCR) testing [4]. It has been observed that RT-PCR also has several vital limitations. The most pertinent of this limitation is that the test is not universally available. To further compound the drawbacks, the turnaround times for this test is currently lengthy and the sensitivities vary. Some studies have even pointed out that that sensitivity of this test is largely insufficient [4]. To mitigate some of the challenges in rapid screening given the large incidence rate of this virus and limited testing facility, radiological imaging complements and supports immensely stratify therapy options for more severe cases of COVID-19.

Radiological imaging equipment, such as X-ray, are more easily accessible to clinicians and also provide huge assistance for diagnosis of COVID-19. CT imaging and Chest radiographs (CXR) are two of the currently used radiological imaging modalities for COVID-19 screening. Lung CT can detect certain characteristic manifestations associated with COVID-19. Several studies [5] [4] have demonstrated that CT is more sensitive to detect COVID-19, with 97-98%, compared to 71% for RT-PCR [4]. CXR might have lesser scope in the first stages of the disease as the changes are not evident on CXR. Studies have shown [6], [7] that CXR may even present normal in early or mild disease, as demonstrated in Figure 1 [8]. CT is hence preferred for early stage screening and is also generally better than X-rays as it enables three dimensional views of the lung.

**Fig. 1.**
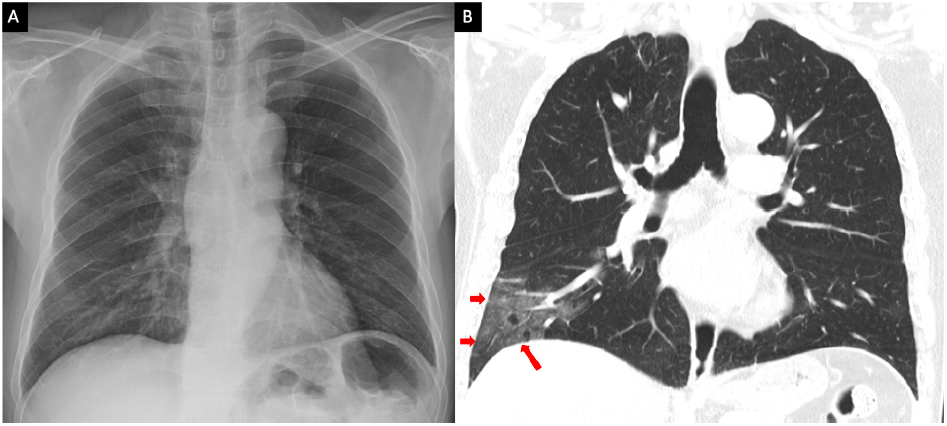
Comparison of chest radiograph (A) and CT thorax coronal image (B). The ground glass opacities in the right lower lobe periphery on the CT (red arrows) are not visible on the chest radiograph, which was taken 1 hour apart from the first study. Image courtesy - Ming-Yen et al [9]

The typical signs of COVID-19 infection observed in CT slices are ground glass opacities (GGO), which occur in the early stages and pulmomary consolidation, which occur in later stages. Detection of these regions in CT slices gives vital information to the clinicians and helps in combating COVID-19. Manual detection is laborious, highly time consuming, tedious and error prone. It has to be pointed out that COVID-19 associated abnormalities, such as ground glass opacities and consolidations, are not characteristic for only COVID-19 but can occur in other forms of pneumonia.

Deep Learning plays a vital role in processing these medical images and correctly diagnosing patients with COVID-19. In regular clinical workflow, while assessing the risks for progression or worsening, the images need to be segmented and quantified. Deep learning based algorithms are able to automatically segment images when trained on manually segmented lesion labels. Several researchers have already established the efficacy of such algorithms on COVID-19 images. One of these early works was DenseUNet proposed by [10] to segment the lesions, lungs and lobes in 3D. They compute percentage of opacity and lung severity scores and report this on entire lung and lobe-wise. The algorithm was trained on 613 manually delineated CT scans (160 COVID-19, 172 viral pneumonia, and 296 ILD). They report Pearson Correlation coefficients between prediction and ground truth above 0.95 for all the four categories. In CovidENet [11] propose a combination of a 2D slice-based and 3D patch-based ensemble architectures, trained on 23423 slices. Their finding was that CovidENet performed equally well as trained radiologists, with a Dice coefficient of 0.7.

For the diagnosis of lung diseases, CT scans have been the preferred modality, and this has therefore been actively utilized in managing COVID-19 [12] [13] [14]. AI in medical imaging has largely aided in automating the diagnosis of COVID-19 from medical images [15] [16]. A detailed review of AI in Diagnosis of COVID-19 has been presented by Shi et al [12]. They broadly group AI based automated assistance for image acquisition, accurate segmentation of organs and infections and for clinical decision making. Under the segmentation approaches they have comprehensively covered nearly all the research that has happened so far in the automated segmentation of lung regions and lesion regions in CT and Xray images.

A variant of inception network was proposed by Wang [16] for classifying COVID-19 from healthy. U-Net++ architecture [17] has been effectively put to use for COVID-19 diagnosis, which worked better than expert radiologists. In the realm of segmentation based methods, [18] and [19] use VB-net [20] to segment of lung and infection regions in CT images. Chaganti et. al [10] use DenseUNet to 3D segment the lung and the lesions.

Fan et al. [21] have reported in their paper the list of public COVID-19 imaging datasets. As mentioned in their paper, there is only one dataset which provides segmentation labels [22]. From this public database [22], we have combined the first dataset of 100 sparsely selected axial CT slices from over 40 patients with a dense set of slices from 9 patients CT scan and use this larger datasets for our studies. A few exemplary slices are demonstrated below to get a visual impression of how the Ground-glass opacity lesions and Consolidation lesions manifests itself in Figure 2.

**Fig. 2.**
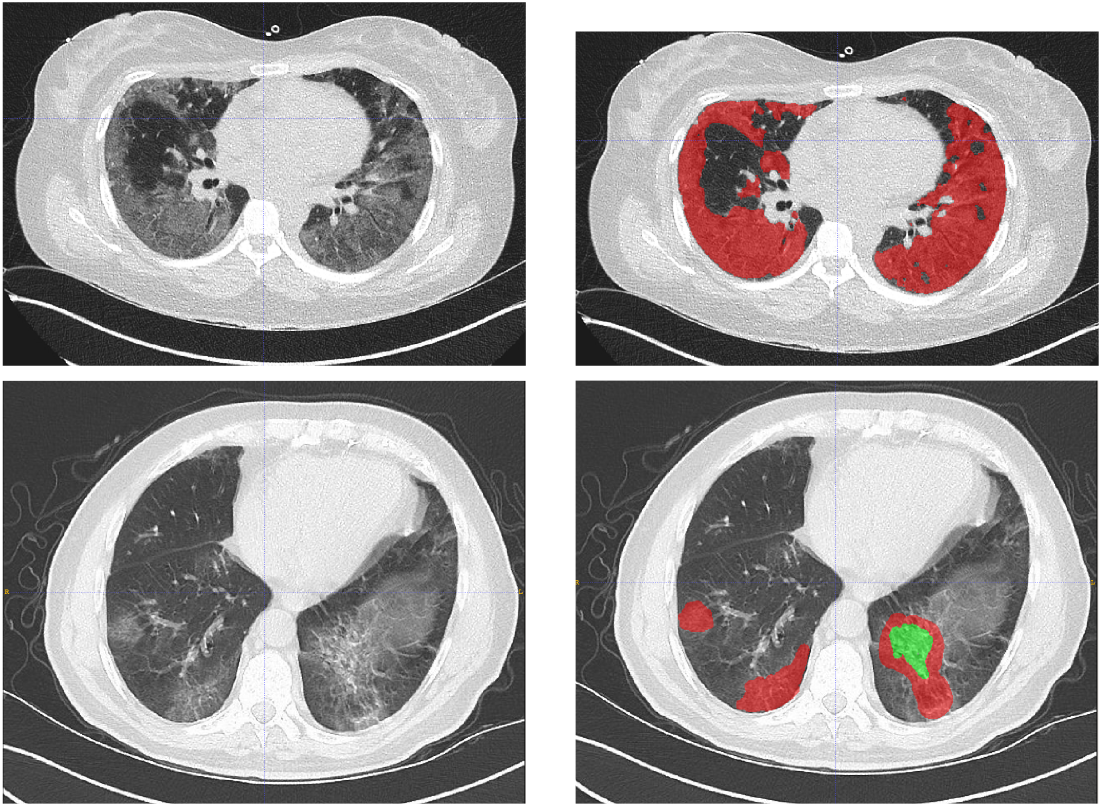
Sample slice from one of the dataset and the corresponding Ground-glass opacity lesion (GGO) marking in first row and GGO and Consolidation lesion marking in second row. Dataset from website [22]

In this paper, we propose Dynamic Deformable Attention Network (DDANet), a novel deep network for COVID-19 infection segmentation in 2D CT slices. Our inspiration for this network is the recent success of self attention mechanisms and sparse deformable convolutions [23]. Attention blocks do not have to be regularly structured, this opens the novel research area that motivates our investigation of spatially-adaptive attention filters. In this work we generalize criss-cross attention [24] for semantic segmentation tasks. We enhance the criss-cross attention and propose a novel deformable attention in which both the attention filter offsets and coefficients are learnt in a continuous, differentiable space. We carried out extensive experiments of our novel algorithm on a large publicly available COVID-19 dataset. Our proposed DDANet achieves very good lesion segmentation and outperforms most cutting-edge segmentation models reported so far on Ground-glass opacity and consolidation lesions. The proposed solution greatly enhances the performance of the baseline U-Net architecture [25]. The baseline U-Net we have employed in our work is from Oktay et al. [25], which has a well-proven strong baseline. Our novel adaptation of the criss-cross attention module is generic and can also be easily plugged into any state-of-art segmentation architecture. These results demonstrate that our proposed DDANet can be effectively used in image segmentation in general and COVID-19 automated image analysis in particular and can greatly aid in clinical workflow handling of these images.

In summary, our main contributions in our work are:

- We propose a novel deformable attention module in which sparse attention filter offsets are learnt in a con-tinous differentiable space and can capture contextual information in an efficient way
- We demonstrate that employing this new deformable attention mechanism within the U-Net architecture [25] [1] achieves superior performance of lung infection segmentation compared to conventional U-Nets or U-Nets with criss-cross attention [24]
- The DDANet reaches state-of-the-art segmentation performance of 73.4% and 61.3% for Ground-Glass-Opacity and Consolidation lesions, on a large publicly available CT COVID-19 infection dataset in a three-fold cross validation on GGO and consolidation labels.

## II. Related work

We discuss two areas of research that are related to our work - Semantic segmentation and Attention mechanism, specifically the criss-cross attention module.

### A. Semantic Segmentation

Semantic segmentation has steadily progressed in the last few years evolving from Fully Convolutional Network (FCN) [26], to the use of dilated convolutions [27] and extensive adaptation of encoder decoder architectures - U-Net [28], Attention U-Net [25] [1], nnU-Net [29], DeepLabv3+ [30], Semantic Prediction Guidance (SPGNet) [31], Discriminative Feature Network (DFN) [32], RefineNet [33] and MultiScale Context Intertwining (MSCI) [34]. To detect objects of various scale, the convolution operator has been enhanced using Deformable Convolution [35] [1] and Scale adaptive convolutions [36]. Graphical models have also been employed effectively for the task of semantic segmentation [37] [27].

Attention models initially gathered a lot of traction after the successful introduction of transformer models in Natural Language Processing (NLP) domain [38]. It has been demonstrated that NLP models perform better when the encoder and decoder are connected through attention blocks.

Attention mechanism have subsequently been utilized in computer vision tasks to capture long-range dependencies. The earlier approaches have tried to augment convolutional models with content-based interactions [30] [39] [1]. The seminal work in attention mechanisms was non-local means [40], which was then followed by self-attention [39]. These have helped achieve better performance on computer vision tasks like image classification and semantic segmentation. Attention-gates have also shown promising results when incorporated into U-Nets for 3D medical segmentation [1]. There have also been successful experiments of building pure selfattention vision models [41].

Non-Local Networks [40] enable full-image context information by utilizing self-attention which helps reference features from any position to perceive the features of all other positions. The drawback of Non-Local network is the large time and space complexity (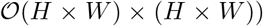) to measure every pixel-pair relation, and also requiring large GPU memory to train such models.

CCNet [24] elegantly solves the complexity issue by using consecutive sparse attention. With two criss-cross attention modules, CCNet captures contextual information from all pixels with far less time and space complexity.

### B. Criss-Cross Attention Module

The criss-cross attention module (CCA) proposed by Huang et al. [24] aggregates contextual information in horizontal and vertical directions for each pixel. The input image **X** is passed through convolutional neural network (CNN) to generate the feature maps **H** of reduced dimension. The CCA module comprises of three convolutional layers applied on **H** ∈ ℝ *^C^*^×^*^H^*^×^*^W^* with 1 _×_ 1 as kernel size.

First, the local representation feature maps H are fed into two convolutional layers in order to obtain two feature maps - query **Q** and key **K** with the same reduced number of feature channels *C*′. By extracting feature vectors at each position u from **Q**, a vector **Q**_u_ ∈ ℝ*^C^*′ is generated. From **K** feature vectors in the same row and column as *u* are collected in **Ω_u_** ∈ ℝ^(^*^H^*^+^*^W^*^−1)×^*^C^*^′^ with elements **Ω_i,u_** ∈ ℝ*^C^*′.

Attention maps **A** ∈ ℝ^(^*^H^*^+^*^W^*^−1)×^*^H^*^×^*^W^* are obtained by applying the affinity operation 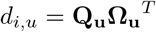 with *d_i,u_* ∈ **D** being the degree of correlation between feature **Q_u_** and 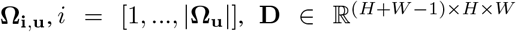 followed by a softmax layer on **D** over the channel dimension.

The third convolutional layer applied on **H** generates Value **V** ∈ ℝ*^C^*^×^*^H^*^×^*^W^* for feature adaption. Therefore, a feature vector **V_u_** ∈ ℝ*^C^* and a set **Φ_u_** ∈ ℝ^(^*^H^*^+^*^W^*^−1)×^*^C^* are extracted at each position *u* in the spatial dimension of **V**.

The contextual information is aggregated by

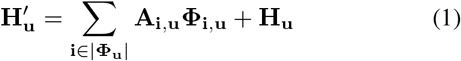

with 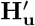 being a feature vector in the module’s output feature maps **H**′ ∈ ℝ*^C^*^×^*^H^*^×^*^W^* at position *u* and **A_i_**, **u** being a scalar value at channel *i* and position *u* in **A**. Finally, the contextual information is weighted with a learnable scalar γ and added to the feature map **H**.

CCNet [24] was shown to enable improvements in computer vision semantic segmentation tasks on Cityscapes, ADE20K datasets. Tang et al. [42] have successfully employed criss cross attention in medical organ segmentation (lung segmentation). In their XLSor paper [42] they used a pretrained ResNet101 replacing the last two down-sampling layers with dilated convolution operation.

The aim of our work is two-fold. First, we evaluate whether criss-cross attention can be employed within a U-Net [1] to improve medical image lesion segmentation for labelled data which is relatively small, a common scenario currently for COVID-19. Second, we incorporate our novel adaptation of this attention model and extend it with a dynamic deformable attention mechanism where the attention filter offsets are learnt in a continuous differentiable space. We strongly believe that the deformable attention module that automatically adapt their layout is an important step to get better insight into the computation mechanism of attention modules. We have discovered in our work that capturing attention from all non-local locations does negatively impact the accuracy of semantic segmentation networks. Capturing only the necessary and essential non-local contextual information in a smart and data driven way yields far more promising segmentation results. We also demonstrate that having the attention offsets learnable enables the network to smartly decide on its own the locations where to obtain non-local attention from for improved results.

## III. Methods

In this section we explain the details of the proposed network architectures. The basic idea of all variants presented is that an attention module is integrated within the U-Net architecture [1] as an extension of the U-Net’s bottleneck in order to capture contextual information from only the necessary and meaningful non-local contextual information in smart and efficient way. Our models utilize the approach of the criss-cross attention module proposed by [24] and modify it to enhance the segmentation performance on COVID-19 datasets.

### A. Network Architecture

The architecture of our model combines the concepts of U-Net [28] and CCNet [24]. A block diagram of the proposed Deformable Attention Net (DDANet) is shown in Figure 3.

**Fig. 3.**
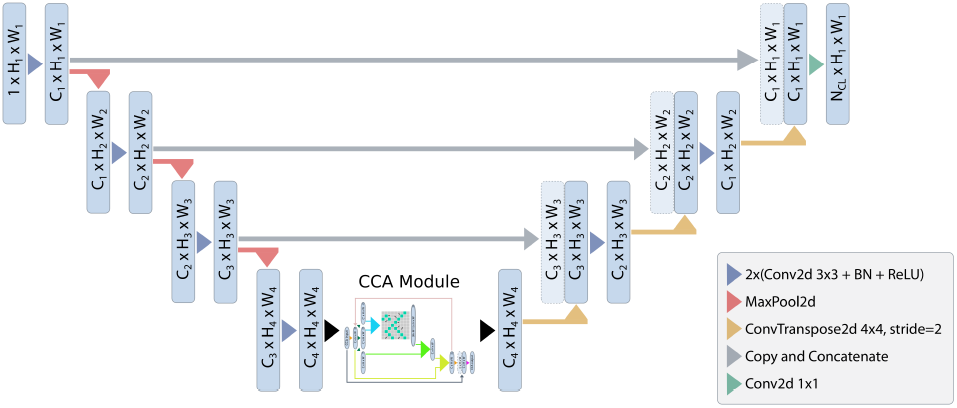
A block diagram of the proposed Deformable Attention Net (DDANet). Input image is progressively filtered and downsampled by factor 2 at each scale in the encoding part. The deformable criss-cross attention is inserted as an extension of the U-Net’s bottleneck in order to capture contextual information from only the necessary and meaningful non-local contextual information in smart and efficient way.

We use a U-Net structure from Oktay et al. [25] [1], adapting it slightly by reducing one downsampling (and corresponding upsampling path), to best process our image dimension (256*256). It consists of three blocks in the downsampling path and three blocks in the upsampling block. Each block consists of 2 × (Batch Normalization - 2D Convolution (kernel size 3 × 3, stride 1, padding 1) - ReLU). The last block consists of a 2D convolution with kernel size 1 × 1. For downsampling, max pooling is applied in the downsampling path to halve the spatial dimension of the feature maps after each block. In the upsampling path ConvTranspose2d is used to double the size of the spatial dimension of the concatenated feature maps. The number of feature channels is increased 1–64–128–256–512 in the downsampling path and decreased again accordingly in the upsampling path. The U-Net’s last layer outputs a number of feature channels matching the number of label classes for semantic segmentation.

The local representation feature maps **H** being output from the U-Net’s last block within the downsampling path serve as input of reduced dimension to the criss-cross module. The attention module is inserted in the bottleneck, as the feature maps are of reduced dimension, and hence the attention maps have smaller, more manageable time and space complexity. In the orginial CCNet [24], the following attention module gathers contextual information in the criss-cross path of each pixel leading to feature maps **H**′. In our proposed novel DDANet, the pattern is dynamic and learnable, and hence a dynamic deformable criss-cross path is used to obtain the attention feature maps **DH**′. These feature maps are again passed through the dynamic deformable attention module again which results in feature maps **DH**″ capturing attention information from the most relavant locations from the whole-image at each of its positions. The contextual features **DH**″ obtained after passing *R* = 2 loops through the attention module are concatenated with the feature maps **X** and merged by a convolutional layer. The resulting feature maps are then passed through the U-Net’s upsampling path.

We implement the following modifications of the criss-cross attention module: Deformable CCA module with *R* = 2 loops, **X** + γ**DH**″

#### Differentiable attention sampling

Consider a classical criss-cross attention operation which gathers non-local information on a feature map of Height *H* and width *W*. The initial shape of the criss-cross pattern is a cross as the orginial CCNet [24] which aggregates contextual information for each pixel in its criss-cross path. We have realized the baseline criss-cross attention by first initializing statically defined locations in a 2D flow field (sampling grid), of size *H* * *W*. The attention filter offsets for the vertical direction is defined as the locations where the x coordinates matches a tensor of length *H* equally spaced points between − 1 and 1. Similarly, the attention filter offsets for the horizontal direction is defined as the locations where the *y* coordinates match tensor of length *W* equally spaced points between −1 and 1. These vertical and horizontal offsets help to compute the attention along a cross pattern at **H** + **W** non-local locations.

To make the attention map differentiable, we compute displacement for the horizontal and vertical offsets. For computing the displacement for each of the horizontal and vertical locations we use *H*+*W* random locations sampled from a standard normal distribution. We distribute these displacement locations smoothly by convolving them three times with gaussian kernel with a kernel size 5. We then use spatial transformer network to sample the attention values from the offset locations coupled with the displacements. To obtain the attention output for inputs on a discrete grid, we use differentiable bilinear interpolation. This makes our attention sampling differentiable and the attention locations are dynamic and deformable.

We realized our dynamic deformable attention mechanism by the differentiable attention sampling described above which deforms the criss-cross pattern. In our deformable attention implementation, we have included **H** + **W** learnable attention offset parameters in our deep neural network definition. These are the learnt displacements for each of the criss-cross locations. The learnt displacement vector (x and y displacement) for each of the criss-cross locations is used to displace the horizontal and veritcal offsets, while sampling the attention maps. For the second recurrence, a second set of different **H** + **W** learnable attention parameters is used for determining the displacements.

We use differentiable bilinear interpolation to differentiably sample the attention values for the query, key and value feature maps from the deformed and dynamically learnt positions of criss-cross offset locations. Hence the attention filter offsets for each of the original criss-cross pattern are learnt in continuous differentiable space. The proposed deformable crisscross attention is depicted in the CCA-Module in Figure 4. As depicted in the figure, the criss-cross pattern is learnt and dynamically deformed to best capture the most relevant nonlocal information.

**Fig. 4.**
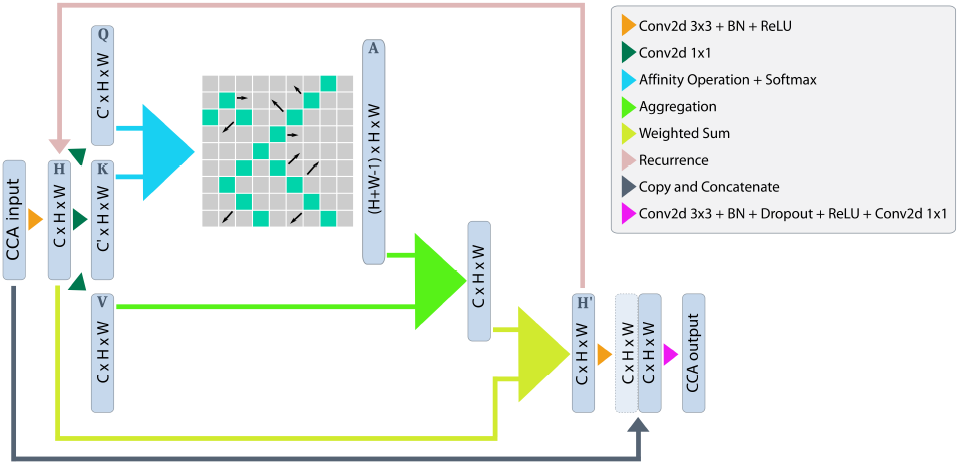
A block diagram of the proposed deformable criss-cross attention module. In our deformable criss-cross, we have the **H** + **W** − 1 learnable attention offset parameters for each of the criss-cross locations. Differentiable bilinear interpolation is used to sample the attention values for the query, key and value feature maps from the learnt positions of deformed criss-cross offset locations.

**Fig. 5.**
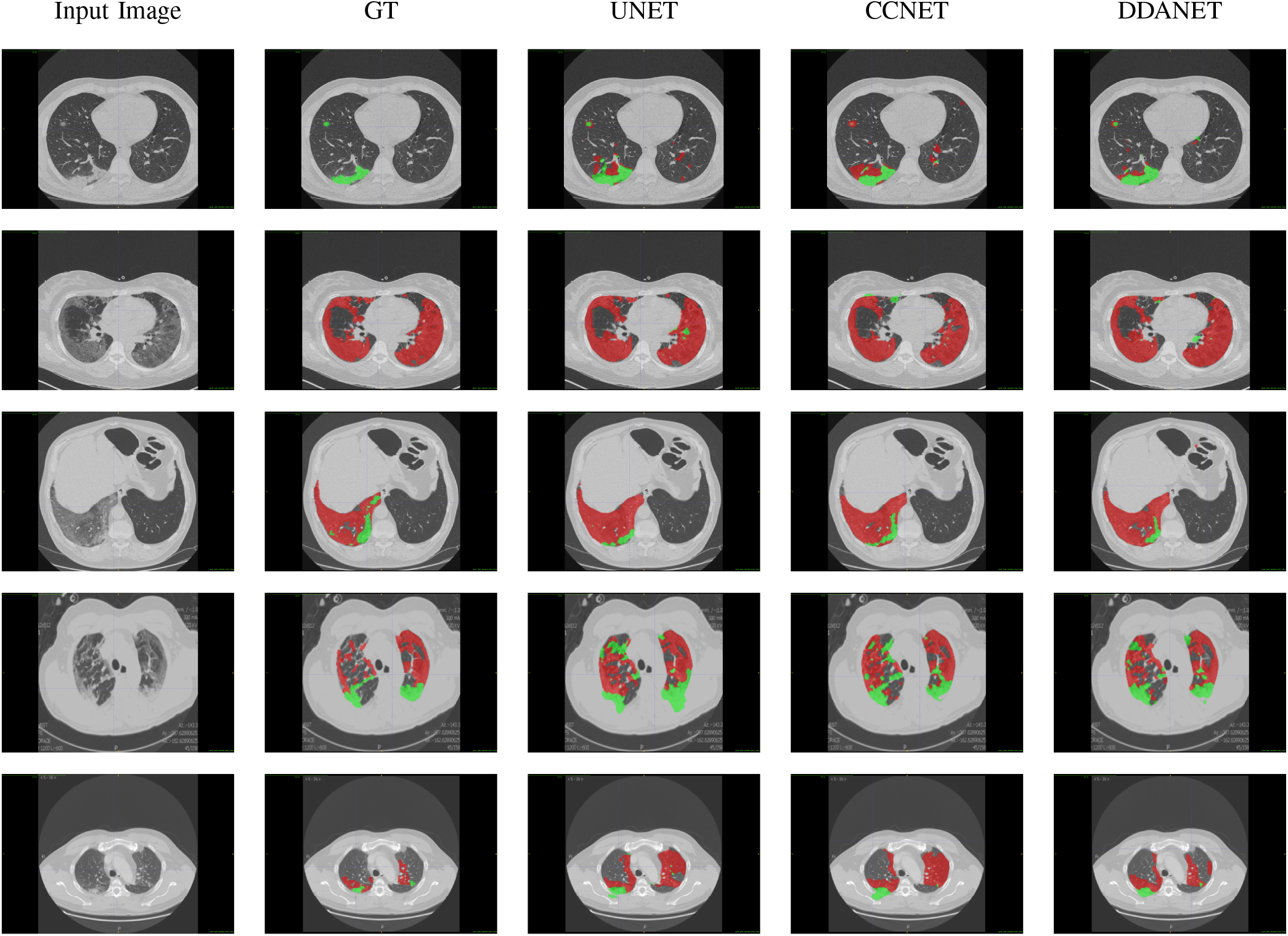
Visual comparison of multi-class lung segmentation results, where the red and green labels indicate the GGO and Consolidation, respectively

The infection class in COVID-19 data is generally under represented as compared to the background class especially in early stages of the disease. This leads to a large class imbalance problem. As found in several studies, Ground-glass opacities generally precede consolidations lesions. This progression of the lesion development in COVID-19 leads to the another scenario of class-imbalance. In some patients only one of the lesions is largely present and the second lesion is highly under-represented (less than 10% of the total infection labels. This also leads to a second category of class-imbalance. To address all of these class-imbalance issues, especially present in COVID-19 lesion segmentation scenarios, we propose to use the inverse class-weighted cross-entropy loss. The weights are computed to be inversely proportional to the square root of class frequency. Given a sample with class label *y*, this inverse class-weighted cross-entropy loss can be expressed as

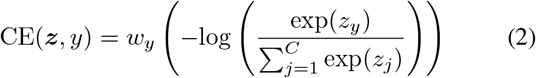

with *C* being the total number of classes and *z* the output from the model for all classes. The weighting factor

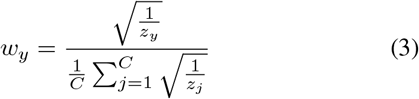

is determined with help of the inverse square root of the number of samples in each label class to address the problem of training from imbalanced data. The training and validation sets also have different distributions, hence we have computed the inverse weighting separately for the train and validation sets. We have also used learning rate finder [43] to find the optimal learning rate, and a 1cycle learning rate policy scheduler, where the maximum learning rate was also determined using the learning rate finder.

## IV. Experimental Setup and Results

We have used the publically available COVID-19 CT segmentation dataset [22]. We have taken the 100 axial CT images from different COVID-19 patients. This first collection of data is from the Italian Society of Medical and Interventional Radiology. We have also utilized the second dataset of axial volumetric CTs of nine patients from Radiopaedia. This second dataset with whole volumes having both positive (373 positive) and negative slices (455 negative slices). We perform experiments with a 3-fold cross validation on this combined dataset consisting of 471 two-dimensional axial lung CT images with segmentations for ground glass opacities (GGO) and consolidation lesions. Each fold comprises data acquired from three different patients plus one third of images from the 100 slice CT stack taken from more than 40 different patients. The CT images are cropped and rescaled to a size of 256 × 256. During training, we perform random affine deformations for data augmentation.

Training is performed for 500 epochs using the Adam optimizer and an initial learning rate of 0.002. We further use a cyclic learning rate with an upper boundary of 0. 005 and a class-weighted cross-entropy loss to address the problem of training from imbalanced data.

For the infection region experiments and multi-class labeling we compared our model with two cutting-edge models: U-Net [25] and Criss-Cross Attention [24]. The number of trainable parameter for the U-Net [25] is 611K. For the U-Net incorporated with the criss cross attention the parameter count is 847K. Our proposed variant of modified CCNet has slightly more parameters at 849K. We have used four widely adopted metrics, *i.e*., Dice similarity coefficient, Sensitivity (Sen.), Specificity (Spec.) and Mean Absolute Error *(MAE)*. If we denote the final prediction as **F_p_** and the object-level segmentation ground-truth as **G**, then the Mean Absolute Error which measures the pixel-wise error between final prediction and ground truth is defined as

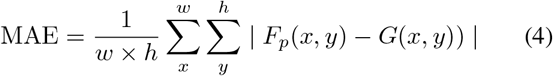

We have adopted a similar approach to Fan et al. [21] and present first the results of our proposed DDANet on detecting lung infections. Our network is trained on multi-class lung infection (GGO and consolidation) and during evaluation we combine these multiple classes into one infection label. We present our 3-fold cross-validation studies results in Table I, which is averaged over multiple runs that we have conducted. We have also included the results from Fan et al. [21] in each of our experiments. It has to be noted that Inf-Net was only trained with the first dataset which is smaller (100 axial slices) and Semi-Inf-Net was trained with pseudo labels from unlabelled CT images. As captured in the Table **I**, our proposed DDANet achieves the best Dice scores in each of the folds. The best Dice score obtained is **0.814** and least mean absolute error (MAE) is **0.0185**. We have also captured the average infection segmentation performance of our network in the same Table **I**. Our proposed DDANet has the best infection segmetation performance in average with the average Dice score of **0.791**). In terms of Dice, our proposed DDANet out-performs the cutting-edge U-Net model [25] by **1.91**% on average infection segmentation.

**TABLE I.** Performance (averaged) of Infection regions on COVID-19 datasets. We have split our data into three folds, and the Results here are averaged over multiple runs for each fold. These are quantitative results of infection regions computed fold-wise and we report 3D Dice-Scores

| Model | Fold | Dice | Sen. | Spec. | MAE |
| --- | --- | --- | --- | --- | --- |
| Inf-Net [21] |  | 0.682 | 0.692 | 0.943 | 0.082 |
| Semi-Inf-Net [21] |  | 0.739 | 0.725 | 0.960 | 0.064 |
| UNET | 0.800<br>0.787<br>0.740 | 0.776 | 0.879<br>0.887<br>0.823 | 0.989<br>0.985<br>0.984 | 0.0208<br>0.0274<br>0.0331 |
| +CCA | 0.809<br>0.798<br>0.735 | 0.781 | 0.876<br>0.888<br>0.850 | 0.990<br>0.986<br>0.981 | 0.0192<br>0.0258<br>0.0357 |
| DDANet | **0.814**<br>**0.808**<br>**0.750** | **0.791** | 0.889<br>0.872<br>0.825 | 0.989<br>0.988<br>0.985 | **0.0185**<br>**0.0240**<br>**0.0318** |

We have also include the infection segmentation performance of our DDANet on each of the Patients in the supplementary materials. In each of the patients, our proposed DDANet had the best Dice score and the minimum MAE. The average across all the patients is captured in Table **II**. In terms of Dice, our DDANet method achieves the best competitive performance of **0.7789** averaged across all the patients. It outperforms the baseline best U-Net model Dice by **3.658**% on infection segmentation.

**TABLE II.** Performance (averaged) on Nine real CTpatient data. These are quantitative results of infection regions computed patient-wise and we report 3D Dice-Scores. The best results are shown in Blue font and the Gain with respect to baseline UNet is shown in Green.

| Model | Dice | Sen. | Spec. | MAE | % Gain |
| --- | --- | --- | --- | --- | --- |
| Inf-Net [21] | 0.579 | 0.87 | 0.974 | 0.047 |  |
| Semi-Inf-Net [21] | 0.597 | 0.865 | 0.977 | 0.033 |  |
| UNET | 0.7515 | 0.8811 | 0.9904 | 0.0149 |  |
| +CCA | 0.7633 | **0.8934** | 0.9908 | 0.0143 | **1.5819** |
| DDANet | **0.7789** | 0.8840 | **0.9915** | **0.0135** | **3.658** |

We have included the fold-wise performance of our DDANet on multi-class labeling in the supplement section. We have captured the average multi-label segmentation performance of our network in Table **III**. We have also compared our results with the results from Inf-Net by Fan et al. [21]. Our baseline U-Net [25] and proposed DDANet has far less trainable parameters at (**611K**) and (**849K**) as compared to **33M** in Inf-Net [21]. Our proposed DDANet has the best multi-label segmetation performance also in average with the best Dice score of **0.734**) for GGO lesions and best Dice score of **0.613**) for Consolidation lesions. Our proposed DDANet has average best dice score of **0.673** for detecting COVID-19 lesions. In terms of Dice, our proposed DDANet out-performs the cutting-edge U-Net model [25] by **4.90**% on average multilabel segmentation. We have increased the trainable parameters in our proposed DDANet only by a neglible amount of 2450 (or 0.3%) in comparison to the original model with criss-cross attention.

**TABLE III.**
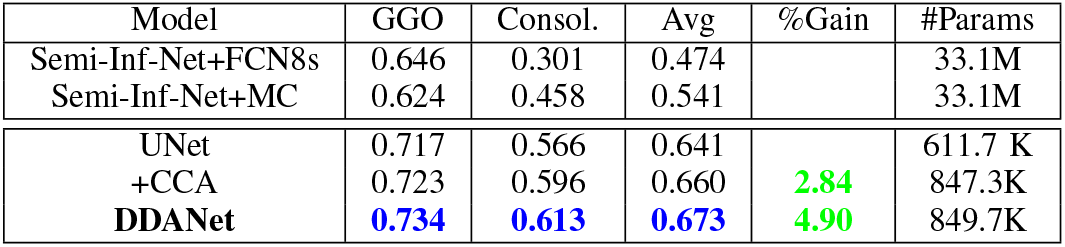
Quantitative results of Ground-Glass opacities and consolidation. The results are averaged across multiple folds and multiple runs. The best results are shown in Blue font.

| Model | GGO | Consol. | Avg | %Gain | #Params |
| --- | --- | --- | --- | --- | --- |
| Semi-Inf-Net+FCN8s | 0.646 | 0.301 | 0.474 |  | 33.1M |
| Semi-Inf-Net+MC | 0.624 | 0.458 | 0.541 |  | 33.1M |
| UNET | 0.717 | 0.566 | 0.641 |  | 611.7 K |
| +CCA | 0.723 | 0.596 | 0.660 | **2.84** | 847.3K |
| DDANet | **0.734** | **0.613** | **0.673** | **4.90** | 849.7K |

We have also captured the multi-label segmentation performance of our DDANet on each of the Patients in the supplementary materials. In terms of Dice, our DDANet method achieves the best competitive performance of **0.702** for GGO lesion and **0.681** for Consolidation lesion averaged across all the patients. In average the proposed DDANet outperforms the baseline best U-Net model Dice by **2.86**% on GGO, **4.73**% on Consolidation and in average **3.52**% on multi-label segmentation. The distribution of the GGO and Consolidation lesions are not even among the different patient scans. Some patients had predominantly only GGO (Patient-8) while other patients had predominantly Consolidation (Patient-3). This skew in distribution impacts the segmentation dice scores significantly, when the lesions are minimally represented in the patients.

## V. Discussion

COVID-19 lesion segmentation is a very challenging problem. One of the major challenge is the regional manifestation of lesions especially in the early stages of the disease, and this can be very hard to get good segmentation in those high class-imbalance scenarios. A similar challenge arises when one of the lesion classes is majorly represented and the other class is highly under-represented which makes it very difficult. This also is a challenging scenario of skewed class-imbalance and gets very hard to get good segmentation in this context as well. The third challenge is the very limited availability of large public datasets, which has been the case until recently. Slowly a number of COVID-19 datasets are made publically available and this scenario could change quite dramatically in the future. This would then enable further research into more compelling algorithms to address this challenging problem.

Our proposed deformable attention is only one of the potential ways to realize learnable attention mechanisms that are smarter elegant and have better performance than earlier proposed criss-cross attention or non-local methods. There are lots of research possibilities to make this even better. There is no requirement or limitation to gather attention from **H+W** locations as we are currently computing. We have currently computed it that way to make it comparable to criss-cross attention. The attention could be gathered from lesser or more locations. One of the next research problems could be to explore what could be the optimal or minimal number of nonlocal attention that needs to be gathered to get the best results. It would also be interesting to establish theoretical upper and lower bounds for number of locations to get non-local attention and its impact on performance. Our work opens up all these and more possible research directions and can be the trigger for more fundamental work on learnable attention mechanisms.

## VI. Conclusion

In this paper, we have proposed a novel adaptation to the criss-cross attention module with deformable criss-cross attention. This has been incorporated into the U-Net framework (DDANet) to improve the segmentation of lesion regions in COVID-19 CT scans. Our extensive experiments have demonstrated that both adapting the U-Net with a straightforward incorporation of the CCNet module and also extending this CCNet with multilpe recurrent application does not yield substantial improvements in segmentation quality. Our novel solution and smart combination of adapted dynamic deformable spatial attention have shown to be a working combination yielding superior and promising results. This solution has immense potential in better aiding clinicians with state-of-art infection segmentation models. For our future studies, we plan to apply explore its adaptation in ResNet like architectures for 2D and once more labelled 3D scans become available the module can easily be adapted to 3D V-Net architectures. We will make our source-code and trained models publicly available.

## Data Availability

The data used is public data

## Supplementary Materials

### I. Introduction

The Supplementary Material section provides the following further details of the main paper topics:

1. Enlarged depiction of the block diagram of our proposed Deformable Attention Net (DDANet) is shown in Figure 1;
2. Enlarged depiction of the block diagram of the proposed deformable criss-cross attention module is shown in Figure 2;
3. One Exemplary Slice Showing Difference between UNet+CCNet (state-of-art) and Proposed DDANet.
4. The infection segmentation performance of our DDANet on each of the Patients in Table **I**;
5. The multi-label segmentation (GGO and Consolidation) performance of our DDANet on each of the Patients through 3D dice scores in Table **II**;
6. The multi-label segmentation (GGO and consolidation), on each of the folds through 3D dice scores in Table **III**. The results are shown across three folds.

We first present the expanded view of the block diagram of the proposed Deformable Attention Net (DDANet) is shown in Figure **1**. The input image is progressively filtered by two consecutive convolution blocks. The number of activation maps or feature channels is increased in the second convolution block. The number of feature channels is progressively increased 1 – 64 – 128 – 256 – 512 in the downsampling path. This double convolution block is then followed by maxpooling lyer. The maxpool layer downsamples the activation maps by factor 2 at each scale in the encoding part. The deformable criss-cross attention is inserted as an extension of the U-Net’s bottleneck in order to capture contextual information from only the necessary and meaningful non-local contextual information in smart and efficient way.

In the upsampling path ConvTranspose2d is used to double the size of the spatial dimension of the concatenated feature maps. The number of feature channels is decreased 512 – 256 – 128 – 64 – *N_CL_* in the upsampling path. The U-Net’s last layer outputs a number of feature channels matching the number of label classes for semantic segmentation.

We next present the expanded view of the block diagram of the proposed deformable criss-cross attention module in Figure 2. In our deformable criss-cross, we have the **H** + **W** − 1 learnable attention offset parameters for each of the criss-cross locations. Differentiable bilinear interpolation is used to sample the attention values for the query, key and value feature maps from the learnt positions of deformed criss-cross offset locations.

**Fig. 1.**
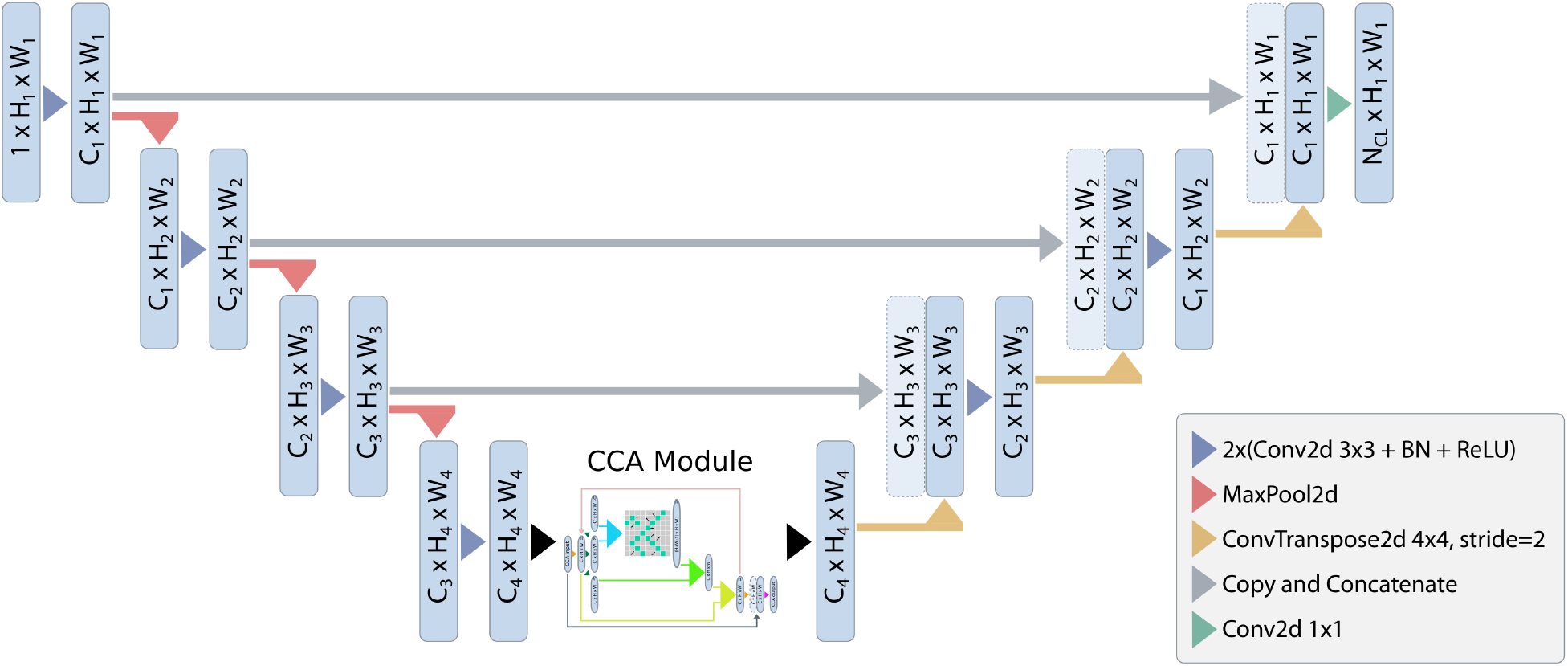
A block diagram of the proposed Deformable Attention Net (DDANet). Input image is progressively filtered and downsampled by factor 2 at each scale in the encoding part. The deformable criss-cross attention is inserted as an extension of the U-Net’s bottleneck in order to capture contextual information from only the necessary and meaningful non-local contextual information in smart and efficient way.

**Fig. 2.**
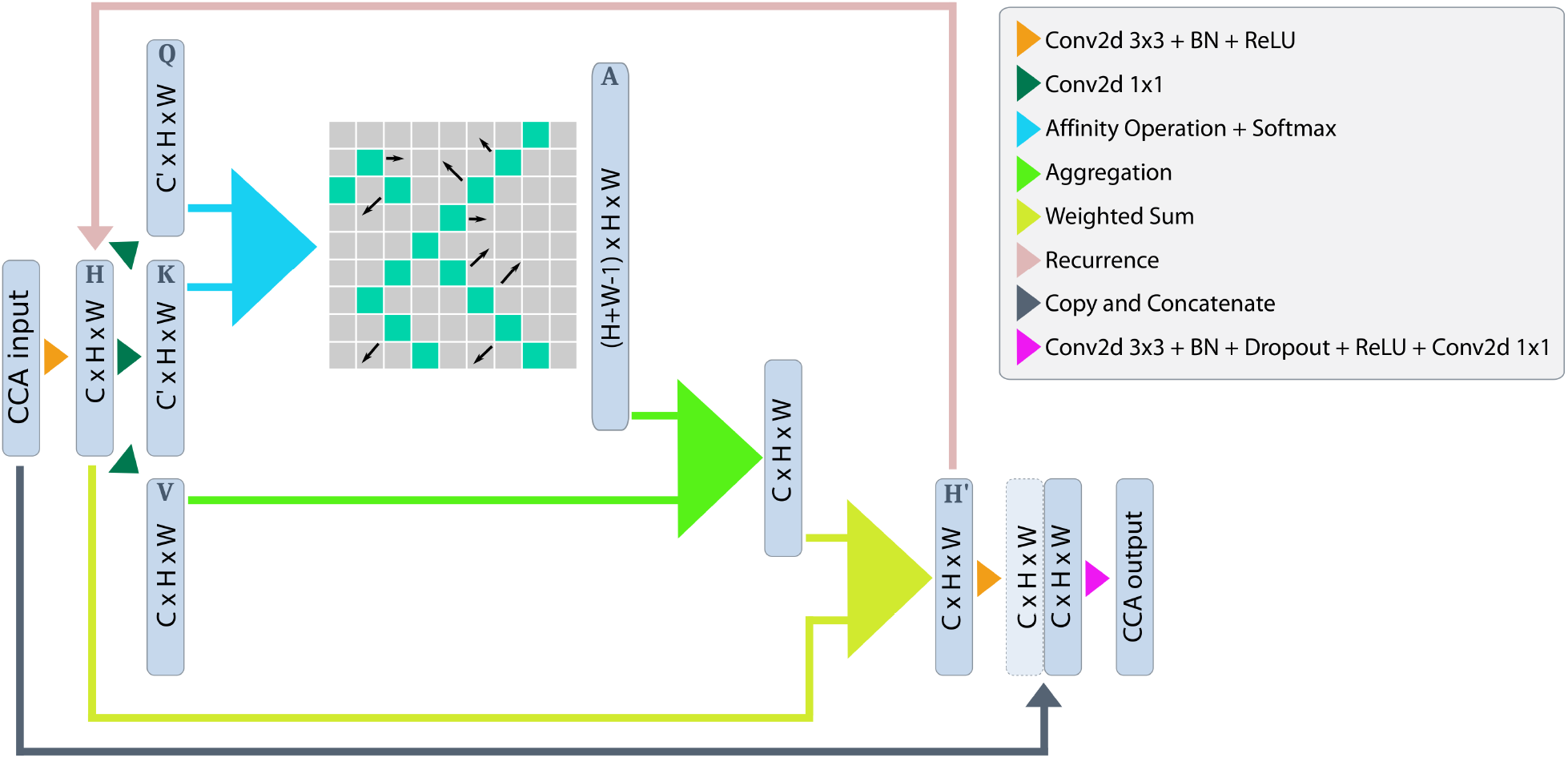
A block diagram of the proposed deformable criss-cross attention module. In our deformable criss-cross, we have the H + W−1 learnable attention offset parameters for each of the criss-cross locations. Differentiable bilinear interpolation is used to sample the attention values for the query, key and value feature maps from the learnt positions of deformed criss-cross offset locations.

**Fig. 3.**
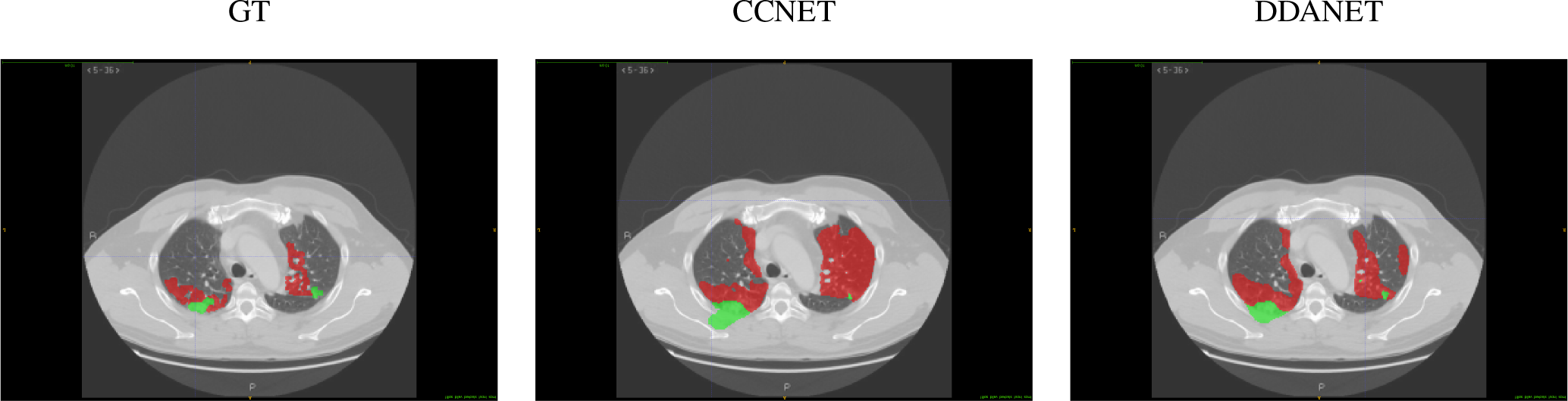
One exemplary slice showing difference between UNet+CCNet (state-of-art) and Proposed DDANet. As is clearly evident UNet+CCNet segmentations leaks into the background when the contrast between structures is smaller and hence it generate spurious segmentations whereas our proposed DDANet has lesser of such leaky effects and has superior performance.

In the Figure 3, we demonstrate one exemplary slice showing Difference between UNet+CCNet (state-of-art) and Proposed DDANet. As is clearly evident UNet+CCNet segmentations leaks into the background when the contrast between structures is smaller and hence it generate spurious segmentations whereas our proposed DDANet has lesser of such leaky effects and has superior performance.

**TABLE I.**
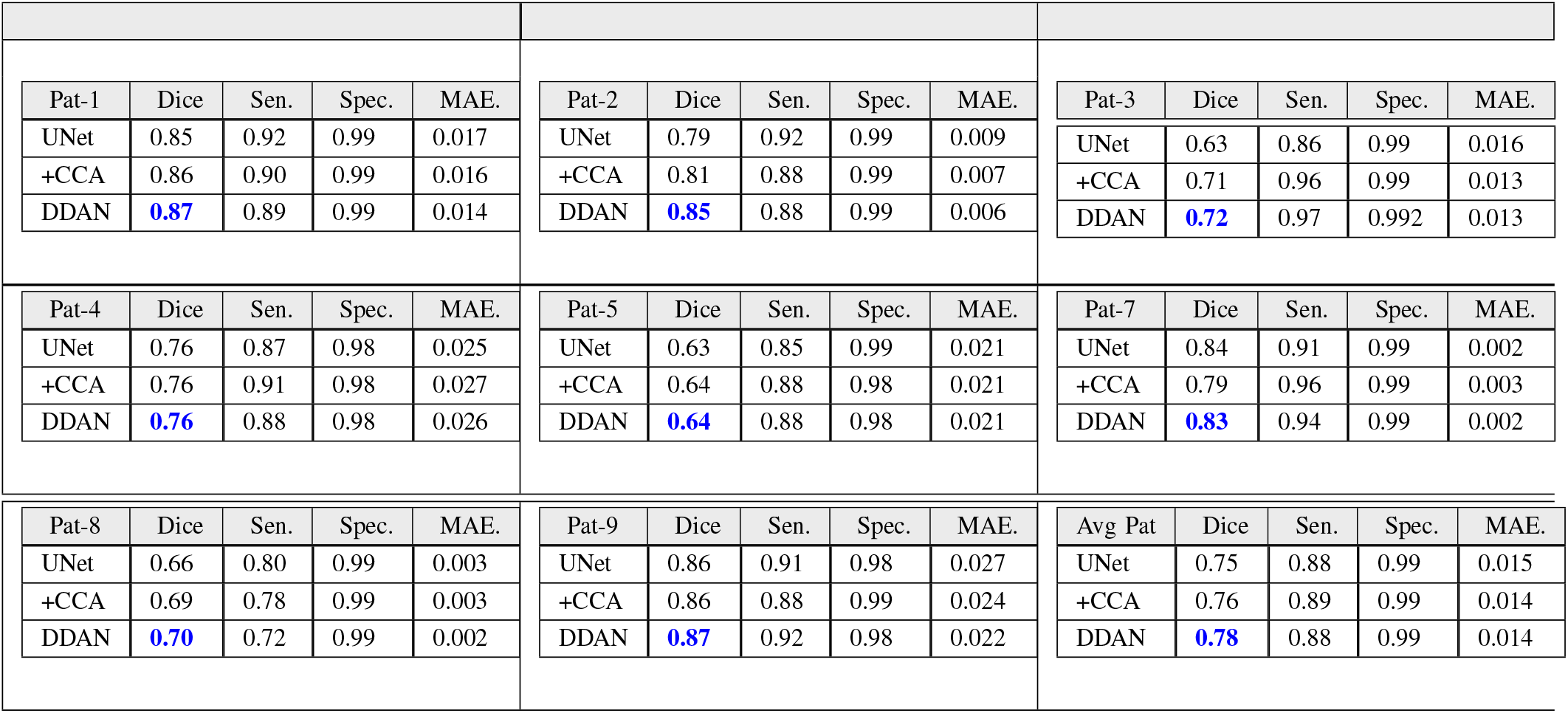
Performance on Nine real CTpatient data. These are quantitative results of infection regions computed patient-wise and we report 3D Dice-Scores

We have also captured the infection segmentation performance of our DDANet on each of the Patients in Table **I**. We have skipped using one Patient (Patient-6) from the dataset, as that had only one slice with infection and only 85 voxels of infection marked in that slice against the total 167M voxels. In each of the patients, our proposed DDANet is having the best Dice score and the minimum MAE. In terms of Dice, our DDANet method achieves the best competitive performance of **0.78** and MAE of **0.014** for Infection segmentation averaged across all the patients.

**TABLE II.**
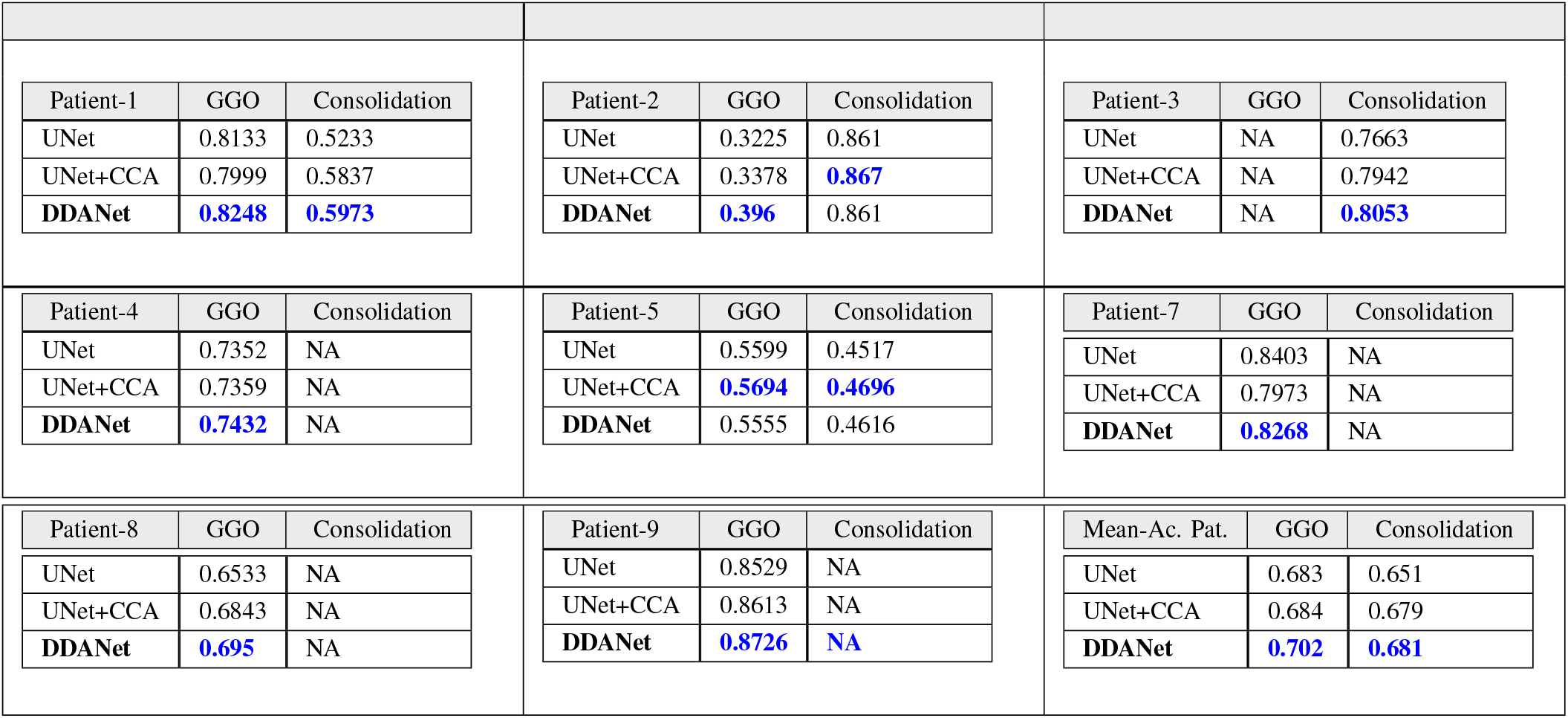
Performance on Nine real CTpatient data. These are quantitative results of multi-label regions computed patient-wise and we report 3D Dice-Scores

We have also captured the multi-label segmentation performance of our DDANet on each of the Patients through 3D dice scores in Table **II**. We have again skipped Patient-6 (due to low lesion representation). The average across all the patients is also captured in the same table in the last block. In six out of the eight patients, our proposed DDANet had the best Dice score for both GGO and Consolidation lesion. In terms of Dice, our DDANet method achieves the best competitive performance of **0.702** for GGO lesion and **0.681** for Consolidation lesion averaged across all the patients.

In average the proposed DDANet outperforms the baseline best UNet model Dice by **2.86**% on GGO, **4.73**% on Consolidation and in average **3.52**% on multi-label segmentation. The distribution of the GGO and Consolidation lesions are not even among the different patient scans. Some patients had predominantly only GGO (Patient-8) while other patients had predominantly Consolidation (Patient-3). This skew in distribution impacts the segmentation dice scores significantly, when the lesions are minimally represented in the patients. We have not taken into consideration those labels in some of the patients when the representation is lower than 10% of the overall lesion distribution as the dice scores gets impacted due to this skewed distribution.

**TABLE III.** Quantitative results of Ground-Glass opacities and consolidation. The results are shown across three folds and averaged over multiple runs. The best results are shown in Blue font and the Gain with respect to baseline UNet is shown in Green.

| Model | Fold | GGO | %Gain | Consol. | %Gain |
| --- | --- | --- | --- | --- | --- |
| Semi-Inf-Net+FCN8s |  | 0.646 |  | 0.301 |  |
| Semi-Inf-Net+MC |  | 0.624 |  | 0.458 |  |
| UNET | fold0 | 0.7687 |  | 0.6799 |  |
|  | fold1 | 0.7225 |  | 0.5699 |  |
|  | fold2 | 0.659 |  | 0.4485 |  |
| +CCA | fold0 | 0.7809 |  | 0.7153 |  |
|  | fold1 | 0.7254 | 0.89 | 0.6055 | 5.3 |
|  | fold2 | 0.6631 |  | 0.4676 |  |
| DDANet | fold0 | **0.787** |  | **0.733** |  |
|  | fold1 | **0.738** | **2.38** | **0.6085** | **8.22** |
|  | fold2 | **0.675** |  | **0.4967** |  |

We have capture the performance of our DDANet on multi-class labeling. We present our 3-fold cross-validation studies results in Table **III**, which is averaged over multiple runs that we have conducted. We have also included the results from Fan et al. [1] in each of our experiments. As captured in the Table **III**, our proposed DDANet achieves the best Dice scores in each of the folds. The Best Dice score achieved for GGO is **0.787** and best Dice score for Consolidation is **0.733**. Our proposed DDANet outperforms the cutting-edge UNet model, in terms of Dice, by **2.38**% in GGO lesion and **8.22**% in Consolidation lesion segmentation in average. Our proposed deformable criss-cross attention is able to segment GGO and consolidation lesions far better than the state-of-art models or baseline UNet models.

